# Over a Billion More Dark Morning School Commutes for K-12 Children under the Sunshine Protection Act

**DOI:** 10.64898/2026.08.24.26361097

**Authors:** Guadalupe Rodríguez Ferrante, Nirekh S. Dasika, Alicia Nam, Jessie Lu, Navneet Tumber, Elena Kully-Rivera, Viridian Klei, Daniel Zhang, Micaela E. Romero, Horacio O. de la Iglesia

**Affiliations:** Department of Biology, University of Washington, Seattle, Washington, USA

**Keywords:** School start times, Daylight saving time, Pedestrian safety, Circadian misalignment, Child and adolescent health

## Abstract

The U.S. House’s approval of the Sunshine Protection Act has revived the debate over permanent daylight saving time (DST) versus permanent standard time (ST). Health and sleep organizations favor permanent ST because it benefits health, especially for children with rigid school schedules. Further, permanent DST would push school start times to before sunrise in many regions, leading to dark-morning commutes. However, the safety consequences of this shift remain unquantified. Using real school start times for 14 states that have enacted permanent DST legislation, together with local sunrise time, we counted the school days on which students must leave home before sunrise under permanent ST, the current system, and permanent DST. In Washington State, where schools start on average at 08:27, neither permanent ST nor the current system requires any pre-sunrise departure, whereas permanent DST would for most of the winter. Using real school start-time data, permanent DST would add about 35 million child-days of pre-sunrise travel in Washington alone relative to the current system, with similar patterns across the other 13 states. Extrapolated to all U.S. public schools and assuming an 8:00 departure, permanent DST would generate more than 2 billion additional dark-morning commutes each year relative to the current system. Finally, analyzing Seattle traffic collisions, we found that the odds that a crash involved a pedestrian were 143% higher on dark mornings (adjusted odds ratio 2.4). Permanent DST would therefore expose many more children, on many more days, to elevated pedestrian-crash risk, evidence that deserves consideration as the United States chooses a time standard.

## Introduction

The recent approval of the Sunshine Protection Act by the U.S. House has reignited the debate on the benefits of permanent daylight saving time (DST) vs. permanent standard time (ST). Human health and safety have been central to this debate. Health, sleep, and scientific organizations have endorsed permanent ST based on the beneficial health effects of aligning solar, biological and social time (1). This alignment is particularly important for children and adolescents. DST delays sunrise by an hour and forces them, particularly adolescents, to wake up during their biological night to comply with rather inflexible school schedules. This mismatch between internal time and socially imposed time leads to unhealthy sleep habits and decreased performance (2, 3).

The safety issue, focused mainly on predicted injuries and deaths as a consequence of car accidents, is still a matter of debate. The predicted safety benefits of one time or the other hinge on whether it is more dangerous to deprive drivers and pedestrians of light during the morning (under DST) or during the evening (under ST). For children attending school, however, the impact of low light levels should focus on the morning, as most children return home under daylight regardless of ST/DST status.

To date, DST has been observed primarily during spring and summer, when sunrise remains relatively early even under DST and when children are largely out of school. Under permanent DST, however, winter school months would bring extremely late sunrise times. Approval of the Sunshine Protection Act by the US Senate would automatically move several states to permanent DST because their respective legislatures have already approved it (4), Washington being among them. Yet the impact of this shift on the safety of school-age children has not been assessed.

Against this backdrop, the objective of this study is to assess how many children and for how many days will be forced into dark morning commutes under permanent DST.

## Results

Permanent DST increases the number of days children and adolescents must leave home before sunrise to reach school (Figure 1). For a school at Washington’s average start time (08:27), and allowing a 20-minute departure buffer for the commute (5), while under permanent standard time or under the current system no day of the school year requires leaving before sunrise (dark commute), under permanent DST, the average school’s students would depart before sunrise for most of the winter (∼November–February; Figure 1a). Averaged across individual students, permanent DST raises dark departures by roughly 30–40 school days per child relative to the current system: from ∼28 days (elementary) to ∼39 days (high school) (Figure 1c). This gradient tracks start time and geographical position: earlier-starting schools accumulate more dark departures, and sunrise happens later in the westernmost parts of the state (Figure 1b).

**Figure 1.**
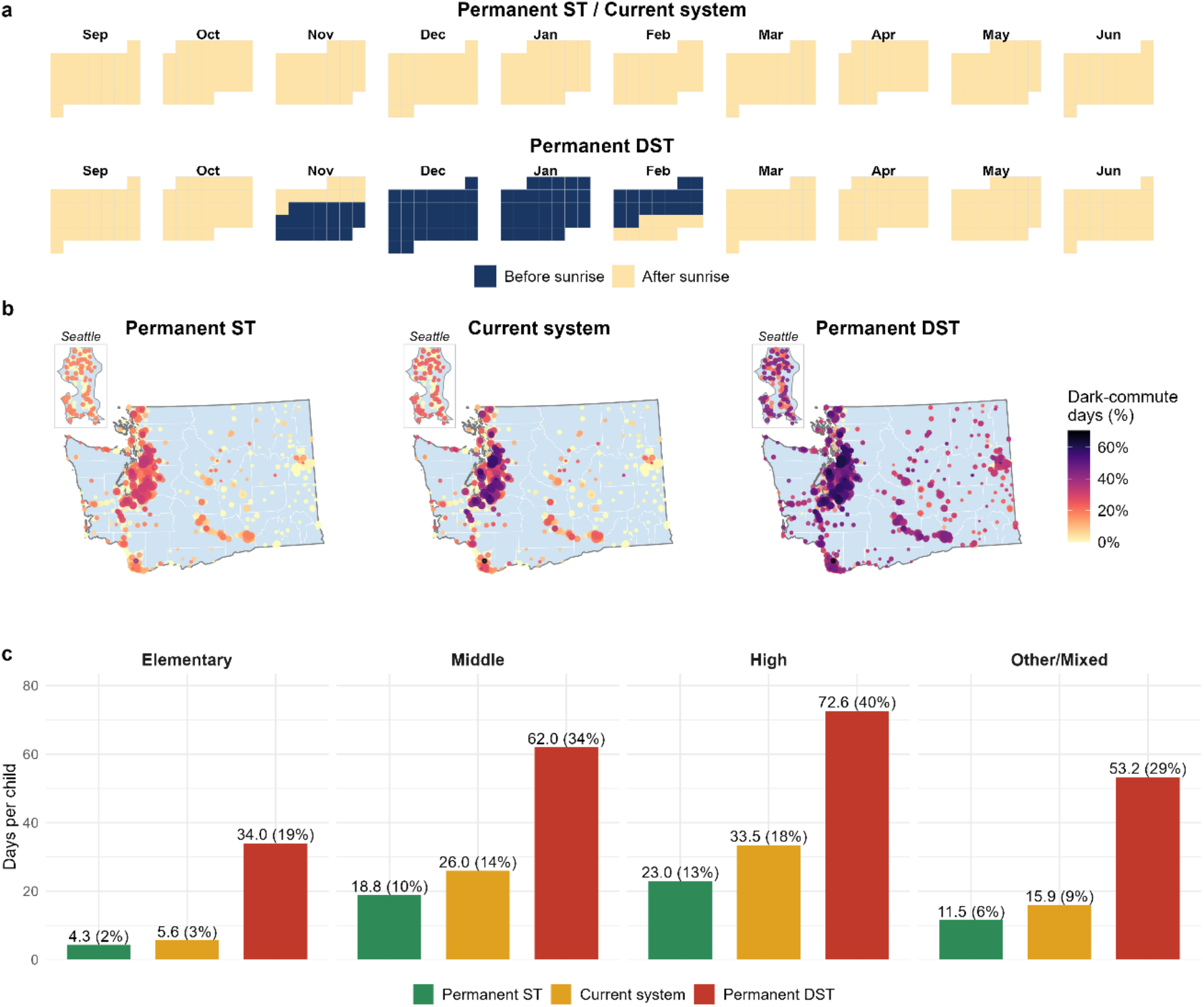
Effect of permanent Daylight Saving Time (DST) on darkness during morning school commutes in Washington State. For all calculations we assume a departure time of school start time minus 20 minutes. (a) School-year calendar showing days on which the weighted average Washington public school’s commute departure time falls before sunrise, under the current clock system (or permanent ST) versus permanent DST. (b) Percentage of dark-commute school days for Washington public schools under permanent ST, the current system, and permanent DST. Point size is proportional to enrollment. Inset: Seattle. (c) Mean dark-commute days per child (raw days, with the percentage of instructional days in parentheses), by school level and clock regime.

Statewide, high schools tend to start earliest and are hit hardest; in contrast, in the city of Seattle, the ordering inverts: elementary schools start earlier (08:02 vs. 08:50 for middle and high) and their students leave before sunrise on most days under permanent DST (≈58 days compared to ≈12–17 days for other school levels).

This carries implications for child safety. Using a 20-year public record of traffic collisions in Seattle (6), restricted to school-year weekdays in the 07:00–08:30 departure window, we found that although the total number of crashes per morning does not differ between dark and light mornings, crashes involving pedestrians are more frequent on dark mornings (0.17 vs 0.10 per morning; rate ratio 1.7). Adjusting for clock hour and weather, the odds that a crash involves a pedestrian are 143% higher on dark mornings than on light ones (adjusted OR = 2.4). Thus, requiring students to travel before sunrise plausibly exposes them to conditions associated with substantially higher pedestrian-crash risk.

In Washington State alone and using real start time data for each school, permanent DST would increase exposure by ≈35 million child-days of pre-sunrise school travel relative to the current system, and ≈41M relative to a permanent ST schedule. This is not specific to a single northern state: the same pattern appears across 13 other states that have already approved permanent DST locally, and that span a wide range of latitudes and time-zone positions (Table 1).

**Table 1.** Dark-morning school commutes by state under three clock regimes. Fourteen of the 19 states that have enacted permanent daylight saving time. Child-days = enrollment × days a student departs before sunrise (start time − 20 min). Students and child-days in millions.

| State | Average SST | N students included | % eligible students included | Child-days departing to school before Sunrise |  |  |
| --- | --- | --- | --- | --- | --- | --- |
|  |  |  |  | Permanent ST | Current System | Permanent DST |
| Washington | 8:27 | 1.11 | 94.4 | 14.01 | 19.74 | 54.96 |
| Oregon | 8:21 | 0.51 | 98.3 | 5.78 | 8.40 | 29.31 |
| Idaho | 8:12 | 0.30 | 100 | 9.10 | 15.22 | 25.42 |
| Utah | 8:21 | 0.66 | 99.1 | 10.11 | 17.99 | 44.14 |
| Colorado | 8:09 | 0.82 | 97.2 | 2.12 | 4.35 | 39.71 |
| Wyoming | 8:11 | 0.09 | 86 | 0.66 | 1.00 | 4.70 |
| Oklahoma | 8:06 | 0.65 | 99.9 | 7.54 | 17.80 | 50.81 |
| Louisiana | 7:48 | 0.66 | 89.3 | 3.62 | 12.23 | 39.54 |
| Tennessee | 7:59 | 0.98 | 95.4 | 8.55 | 18.23 | 56.89 |
| Mississippi | 7:50 | 0.44 | 82 | 0.31 | 1.99 | 20.15 |
| Alabama | 7:52 | 0.75 | 85.9 | 0.65 | 2.00 | 27.69 |
| Georgia | 8:07 | 1.71 | 99.1 | 32.39 | 74.70 | 146.01 |
| Maine | 8:06 | 0.16 | 97.9 | 0.42 | 0.62 | 6.68 |
| Delaware | 8:07 | 0.14 | 99.2 | 1.78 | 3.23 | 7.49 |
| TOTAL (all included states) | 8:08 | 8.99 | 95.2 | 97.02 | 197.51 | 553.50 |

Extrapolated to all U.S. public schools and assuming a fixed 08:00 departure from home, permanent DST would impose 2.14 billion additional dark morning commutes each year relative to the current system.

## Discussion

In this study, we show that under current school start times, adopting permanent DST would substantially increase the number of children who must leave home before sunrise to attend school, not only in a northern state such as Washington, but across several states nationwide. It has recently been argued that permanent DST would be beneficial because it reduces the likelihood of vehicle-deer collisions (7). However, as we show here and consistent with prior work, pedestrian-involved crashes are more frequent during dark mornings than during well-lit ones, meaning that adopting permanent DST would, in turn, expose many more children and for many more days per year to greater traffic accident risk. This risk could be further increased by the fact that the number of vehicle accidents involving high school-age drivers is higher when they commute earlier in the morning (8, 9).

A related consideration is that, over the past few decades, school districts throughout the country have made an effort to delay secondary school start times (10), driven by mounting evidence that early start times are detrimental to adolescents because of the misalignment between the social and solar clock that they produce. Adopting permanent DST effectively advances school start times relative to the solar clock without changing the clock time itself, reversing the positive impacts of delayed school start times, and negatively impacting children’s sleep, physical and mental health, and academic performance.

Beyond their limitations, including that our traffic accident data come from Seattle only and do not distinguish pedestrian age, our results are grounded in real school start times and show that a substantial share of US youth would be affected by the adoption of permanent DST. Altogether, adopting permanent DST would negatively impact youth sleep, health, and performance and, most critically, their safety. As policymakers weigh which time standard the United States should adopt, this evidence deserves a seat at that table.

## Materials and Methods

This study used only publicly available, de-identified data and did not involve human participants. Extended Methods are provided in the Supplementary Information. School enrollment, location, and grade level were obtained from the NCES Elementary/Secondary Information System (ELSi), and each school’s regular instructional start time across 14 U.S. states was collected manually. A nationwide figure was approximated by applying an 08:00 reference departure time to all U.S. public schools. Seattle traffic-collision records (6) were used to relate morning darkness to pedestrian involvement. See Extended Methods (SI Appendix) for details.

## Data Availability

Code and data underlying this study are maintained in a private GitHub repository, which will be made public and archived on Zenodo with a citable DOI upon publication of the manuscript. Prior to that, data are available from the corresponding author upon reasonable request.

https://data-seattlecitygis.opendata.arcgis.com/datasets/SeattleCityGIS::sdot-collisions-all-years/about

## Acknowledgments

We thank Esteban Miglietta, Ishan Mehta, Justin W. Kahn, Paolo Almario, Elshaday Fekadie, Jasmine Nguyen, Yuna Yuan, Raymond Lam, Nancy Ta, Selam Babenga, JD Kim and Talia Trammell for their help in data collection.

## Supporting Information

### Supporting Information Text

#### Extended Methods

##### School start times

School demographic and location data were obtained from the publicly available National Center for Education Statistics (NCES) website using the Elementary and Secondary Information System (ELSi) table generator (1). Schools meeting the inclusion criteria were selected within the ELSi interface and the following variables were exported: School Year, County Name, School ID, District Name, School Name, Total Student Enrollment, Latitude and Longitude. The table was exported to a Microsoft Excel spreadsheet, to which start-time and comments columns were added and completed through manual data collection of each school’s regular instructional start time and any scheduling notes. Start times were sought primarily on official district and school websites (bell schedules, calendars, handbooks, parent/student resources) via targeted searches (e.g., “[district] School District Bell Schedules / Start Times / Arrival Times”); district websites were prioritized over search-engine summaries. When a start time could not be found online, the school was contacted by telephone, email and/or through social media, with unresolved cases annotated. For Tennessee and Colorado, an AI assistant (Claude, Anthropic) was used to help locate candidate start times, but every value was subsequently verified manually against the district source. When more than one bell time was reported, we used class-start time in preference to the tardy bell and the first bell; optional breakfast start times were not considered. Because these choices favor the latest plausible arrival time, our estimates are conservative and likely underestimate the effect of permanent DST. Schools with multiple daily schedules were summarized as the day-weighted average start time across the school week. Enrollment and coordinates correspond to 2024–25. Start times reflect the latest school start time available, in most cases 2025–26 or 2026-27 schedules.

We compiled data for 14 states — Washington, Oregon, Utah, Colorado, Wyoming, Idaho, Oklahoma, Louisiana, Tennessee, Mississippi, Alabama, Georgia, Maine, and Delaware — representing 14 of the 19 U.S. states that have enacted legislation to adopt permanent DST. Inclusion was restricted to in-person schools primarily serving students from elementary through high school. We excluded schools that were virtual/non-in-person schools, preschools, schools with no reported enrollment or that were closed, juvenile detention centers, reengagement programs, technical/adult-serving schools, supplementary-class programs and schools with highly irregular schedules (e.g., alternating morning/afternoon). Special-needs and technical high schools were included when they served adolescents on a consistent schedule. Across the included states and among eligible schools, we located a usable start time for 93% of enrolled children (91% of schools), with a small number further excluded for invalid coordinates, yielding the analytic sample. Specifically, of the total 18,549 schools in the original database, 1,376 were excluded because they met one of the exclusion criteria, and 1,080 because we did not find any start time; thus, we included 16,083 schools serving 8.5M children.

#### Sunrise and clock regimes

Sunrise at each school was computed with suncalc (2) as the upper solar limb at the horizon (altitude −0.833°, including atmospheric refraction), and expressed as clock time under three policies: permanent standard time, the current system, and permanent daylight saving time. For each school and instructional day, we recorded whether departure time (start time minus a 20-min commute) preceded sunrise and summed over enrollment to obtain child-days of pre-sunrise school travel. Each school’s clock offset was set by its time zone: the current system adds one hour during daylight saving months, permanent DST adds one hour year-round, and permanent standard time uses the standard offset throughout. Time zones were assigned per school; for the nationwide computation, coordinates were mapped to IANA time zones with the lutz package (3).

##### Dark-exposure metrics

The instructional calendar comprised school-year weekdays (September–June) minus public holidays and typical breaks (≈183 school days). For each school and instructional day, the start time and the departure time (start – commute and buffer) were compared with the school’s sunrise clock time under each regime. Events preceding sunrise were counted as “dark,” and per-school counts were summed over enrollment to yield child-days of pre-sunrise travel. The commute buffer was 20 min in the main analysis (mean U.S. student travel time ≈18 min (4) plus a 2 min buffer to get to their class). For Washington State school level was assigned based on the school’s name: 1,092 elementary, 373 middle, 366 high, and 184 other/mixed-level schools.

##### Nationwide approximation

Because per-school start times are unavailable nationally, we approximated the national impact by applying the same computation to all U.S. public schools (NCES coordinates and enrollment; time zones via *lutz*) under a fixed 08:00 reference departure time to go to school, reporting the child-days for which departure precedes sunrise under each regime. To bound sensitivity to this assumption, we repeated the computation using departure times of 07:45, 8:00 and 08:15. Compared with the current system, permanent DST would increase pre-sunrise commute time by 2.63 billion, 2.14 billion, and 1.39 billion child-days, respectively. This calculation is intended only to dimension the national scale and is not a substitute for per-school data.

##### Statistical analysis

All inferential analysis concerns the Seattle collision data as dark-exposure quantities are deterministic. Analyses were restricted to school-year weekdays in the 07:00–08:30 window. Darkness was defined by solar altitude (−0.833°) at each collision. The association between darkness and pedestrian involvement was estimated by logistic regression (pedestrian-involved ∼ dark + clock hour + weather; with clock hour included as a factor and weather coded as “Clear”, “Overcast”, “Raining” or “Other”), reporting the adjusted odds ratio (95% CI) and a crude odds ratio (Fisher’s exact test). Total crashes per morning were compared between dark and light mornings.

## Notes

### Competing Interest Statement

The authors have declared no competing interest.

